# Elevated pulse pressure and risk of chronic kidney disease by hypertension status: A longitudinal study in Japanese adults

**DOI:** 10.1101/2025.03.27.25324763

**Authors:** Yukari Okawa, Toshiharu Mitsuhashi

## Abstract

**Introduction:** Little is known about the relationship between pulse pressure (PP) and incident chronic kidney disease (CKD) in Asian populations, particularly when analyzed separately by hypertension status.

**Aim:** This study aimed to assess the association between PP and subsequent onset of CKD in Japanese adults.

**Methods:** This longitudinal study included middle-aged and older Japanese citizens who participated in administrative checkups (1998–2024) conducted by Zentsuji City. The relationship between PP (diastolic blood pressure subtracted from systolic blood pressure) and incident CKD (estimated glomerular filtration rate <60 mL/min/1.73 m^2^) was evaluated by hypertension status, using the Weibull accelerated failure time model. PP was treated as a time-varying variable and categorized into <40 (reference), 40–<60, and ≥60 mmHg. In addition to the crude model, two adjusted models were created to control for potential confounders.

**Results:** Among 15788 participants, 8881 (men: 42.7%) were examined in the study. The mean follow-up time was 6.21 years for non-hypertensive participants and 6.27 years for hypertensive participants. Higher PP was associated with higher rate of CKD incidence regardless of prevalent hypertension. In non-hypertensive participants, PP ≥60 mmHg had a 10% shorter time to CKD onset (95% confidence interval: 3%–16%) compared with PP <40 mmHg. In hypertensive participants, attenuated results were observed, with all 95% confidence intervals crossing the null value.

**Conclusions:** Elevated PP may serve as a useful indicator for CKD development in non-hypertensive Japanese subjects. Regular BP monitoring may assist in developing public health strategies for CKD prevention, especially among non-hypertensive Asian populations.

## Introduction

Pulse pressure (PP) is one of the indicators reflecting arterial stiffness[1]. It can be calculated simply by subtracting diastolic blood pressure (DBP) from systolic blood pressure (SBP). Since blood pressure (BP) measurement is a routine non-invasive test in clinical practice, PP is a useful index for assessing disease risk and prevention. Studies have shown that elevated PP is associated with elevated risks of cardiovascular disease events, stroke, and mortality in older adults, whereas in younger adults, its impact appears paradoxically attenuated[1–6].

PP plays an important role in new-onset chronic kidney disease (CKD). To date, various follow-up studies of adult participants across multiple ethnic groups have investigated the relationship between PP and incident CKD under different conditions. For example, cohort studies have been conducted in Sweden, Japan, and China with different perspectives[2,7,8]. In summary, all three studies indicated that higher baseline PP is associated with incident CKD. However, the studies included different racial populations and used different analytical conditions in middle-aged and older adults. Specifically, the Good Aging in Skåne study in Sweden included Swedish subjects and compared PP 60–<70, 70–<80, ≥80 mmHg versus PP <60 mmHg, the Kansai Healthcare Study in Japan included male Japanese subjects and compared the fourth quartile (PP ≥56 mmHg) versus the first quartile (PP ≤40 mmHg), and the China Health and Retirement Longitudinal Study in China included non-hypertensive Chinese subjects and compared PP ≥60 mmHg versus PP <60 mmHg.

These three studies attempted to mitigate the influence of potential confounders such as age and body mass index (BMI). However, only the China Health and Retirement Longitudinal Study focused on the impact of hypertension status on the results, by specifically investigating non-hypertensive participants[9]. The relationship between PP and hypertension is bidirectional, and hypertension impairs renal function[6,10]. Based on the findings of the National Health and Nutrition Examination Survey in 2011–2014, non-Hispanic Asian people tend to have a slightly lower prevalence of hypertension than non-Hispanic white people[11]. Therefore, to assess the relationship between PP and CKD onset more accurately, it is important to stratify the results by hypertension status to evaluate its impact on PP, not only for both non-hypertensive and hypertensive participants but also within the same racial population to ensure the validity of the results.

The effect of changes in PP over time is not negligible. A meta-analysis on seven cohort studies revealed that more visit-to-visit BP variability was associated with a higher risk of CKD onset[12]. However, previous studies only investigated how baseline PP affects renal function, without considering the time-varying effect of PP throughout the follow-up period[7–9]. Accordingly, the present longitudinal study aimed to determine whether elevated time-varying PP shortens the time to CKD onset in middle-aged and older Japanese adults with and without hypertension.

## Methods

### Data source and study participants

This secondary analysis study included all participants who underwent annual health checkups conducted by Zentsuji City, Kagawa Prefecture, Japan (participation ratio: 30%–40%)[13]. The target populations for these checkups were citizens aged ≥40 years in each fiscal year (FY). In FY1998 and FY1999 only, citizens aged ≥35 years were added to the target populations under the city’s health promotion initiatives for targeting younger citizens. Data for this study were extracted on 1 July 2024 from the same database used in previous studies[14–18].

In this study, all of the participants were Japanese, had BP and renal outcome data, did not have CKD at study entry, and had multiple observations. Missing observations for BMI and liver function tests were removed due to the small proportions of missingness (0.01%).

### Measurements

Each annual health checkup included measurements of height, weight, and BP; blood tests; urinalysis; and a self-reported questionnaire enquiring about medication use, medical history, and lifestyle habits. All measurements were conducted in accordance with the protocol established by the Ministry of Health, Labour and Welfare (MHLW) of Japan[19]. No dietary restrictions were specified for checkups, and all measurements were conducted in both fasting and non-fasting states. Information on the time of the last meal was unavailable for this study. The data used in the study did not contain any medical history based on physician diagnoses.

The MHLW has referred to the handbook published by the Japanese Association Cardiovascular Disease Prevention as a guide for BP measurement[19,20]. According to the handbook, BP should be measured using either the auscultatory method with an electronic (mercury-free) or aneroid sphygmomanometer, or with an upper-arm automatic BP monitor. The MHLW recommends using the average of two BP measurements as the representative value. However, a single reading is considered acceptable when a second measurement is not feasible because of unavoidable circumstances, based on the judgment of the checkup site. (According to questionnaire results from 32 checkup sites in Japan, the majority [68.8%] measured BP twice only when the first reading was elevated, while only 18.8% routinely measured BP twice without exception[21].)

Age in years and biological sex were verified in the city’s database[13]. SBP and DBP (both mmHg) were measured. BMI (kg/m^2^) was calculated by dividing the participants’ weight in kilograms by their height in meters squared. Blood tests included serum creatinine (SCr) test (mg/dL), cholesterol tests (low-density lipoprotein cholesterol [LDL-C], high-density lipoprotein cholesterol [HDL-C], and triglycerides [TG]; all mg/dL), hemoglobin A1c (HbA1c) test (%), and liver enzyme tests (aspartate aminotransferase [AST] and alanine aminotransferase [ALT]; both U/L). SCr was measured by an enzymatic method.

As of FY2013, Japan changed the default HbA1c reporting system to the National Glycohemoglobin Standardization Program (NGSP) from the Japan Diabetes Society (JDS)[22,23]. Therefore, HbA1c_JDS_ was standardized to HbA1c_NGSP_ using the officially certified conversion equation: *HbA1c_NGSP_* = 1.02 x *HbA1c_JDS_* + 0.25[24].

In the questionnaire, smoking status was assessed by asking the participants whether they habitually smoked, which was defined as smoking for at least 6 months or having smoked at least 100 cigarettes in total. Participants were asked about their frequency of alcohol intake (daily/sometimes/seldom or never). The use of antihypertensive agents, insulin, hypoglycemic agents, and antihyperlipidemic agents was asked with a “yes” or “no” response. The MHLW added questions regarding medication use in FY2012 and removed or modified the questions for individuals aged ≥75 years in FY2020, which caused ≥50% of the total observations to be unmeasured[19].

## Variables

### Exposure variable: Pulse pressure

The exposure variable for the study was PP (mmHg), calculated by subtracting DBP from SBP. PP was measured annually and treated as a time-varying variable in the analysis.

Because there is no universally established cutoff value for elevated PP[25], it was empirically categorized into three groups: <40 (reference), 40–<60, and ≥60 mmHg. Following the American College of Cardiology/American Heart Association guideline, normal PP was defined as <40 mmHg, based on the calculation of normal SBP (<120 mmHg) minus normal DBP (<80 mmHg)[11]. The ≥60-mmHg threshold for the highest PP category was adopted from the China Health and Retirement Longitudinal Study[9].

### Outcome variable: Incident chronic kidney disease

The outcome variable was incident CKD. Renal function was assessed by the estimated glomerular filtration rate (eGFR), which was calculated using the three-variable equation for Japanese subjects: *eGFR*(*mL/min/1.73m*^2^) = 194 x *SCr*^-1.094^ x *age*^-0.287^(x 0.739 *if female*)[26,27]. CKD was defined as eGFR <60 mL/min/1.73 m^2^[27].

### Other covariates

Age was treated as a continuous variable. Sex was categorized into male (reference) and female. Overweight or obese was defined as BMI ≥25.0 kg/m^2^ [28]. In accordance with the American College of Cardiology/American Heart Association guideline, hypertension was defined as SBP ≥130 mmHg or DBP ≥80 mmHg[11]. Dyslipidemia was defined based on the Japan Atherosclerosis Society guidelines as LDL-C ≥140 mg/dL, HDL-C <40 mg/dL, or TG ≥150 mg/dL[29]. Self-reported smoking status was dichotomized into non-/ex-smoker (reference) or smoker. Participants were regarded as having diabetes if their HbA1c was ≥6.5%, following the diagnostic thresholds recommended by the American Diabetes Association[30]. Liver function was evaluated using the De Ritis Ratio, calculated as AST divided by ALT[31,32]. Self-reported alcohol intake was categorized as non-/seldom-drinker (reference) or drinker. Information on medication use was excluded from the analysis due to the large proportion of missing data among older participants aged ≥75 years, which may have introduced an age-related bias[19].

### Statistical analysis

All analyses were conducted separately by hypertension status. The dataset was reformatted into a person–period structure, with each period defined by the PP measurement at the start of the interval, which was treated as the baseline PP for that period. The following missing binary variables were filled using a logistic regression imputation method with 40 imputations[33]: self-reported alcohol intake (missing: 31.2%), self-reported smoking status (missing: 30.8%), dyslipidemia (missing: 25.6%), and diabetes (missing: 8.2%)[34]. Participant characteristics during the follow-up period were summarized separately based on prevalent hypertension status, and presented as number of failures, person-years, and incidence rate per 1000 person-years (IR), averaged over 40 imputations if imputed[33]. Mean and standard deviation were shown for continuous variables.

Kaplan–Meier curves were created for the PP categories, separated by hypertension status. Violation of the proportional hazards assumption was confirmed both statistically and graphically[35]. Therefore, the Weibull accelerated failure time model was selected to assess the association between time-varying PP and incident CKD, based on the lower Akaike and Bayesian information criteria values[36,37]. The outcome measure was the time ratio (TR), where TR <1 indicated a shorter time to CKD onset.

In addition to the crude model, Model 1 (adjusted for sex and age) and Model 2 (adjusted for sex, age, overweight or obesity, self-reported alcohol intake, self-reported smoking status, dyslipidemia, diabetes, and AST/ALT ratio) were created, stratified by hypertension status. When interaction effects were observed between the time-varying PP categories and the following variables, a multiplicative term was added to the model to improve the model stability: age, overweight or obesity, self-reported alcohol intake, self-reported smoking status, dyslipidemia, diabetes, and AST/ALT ratio. **Figure 1** shows the causal directed acyclic graph assumed in this study. Because SCr or eGFR at study entry acts as a collider between the exposure and outcome variables, adjusting for it would introduce collider bias and potentially lead to a spurious, non-causal association[38,39]. Therefore, we did not adjust for SCr or eGFR at study entry in this study.

**Figure 1.**
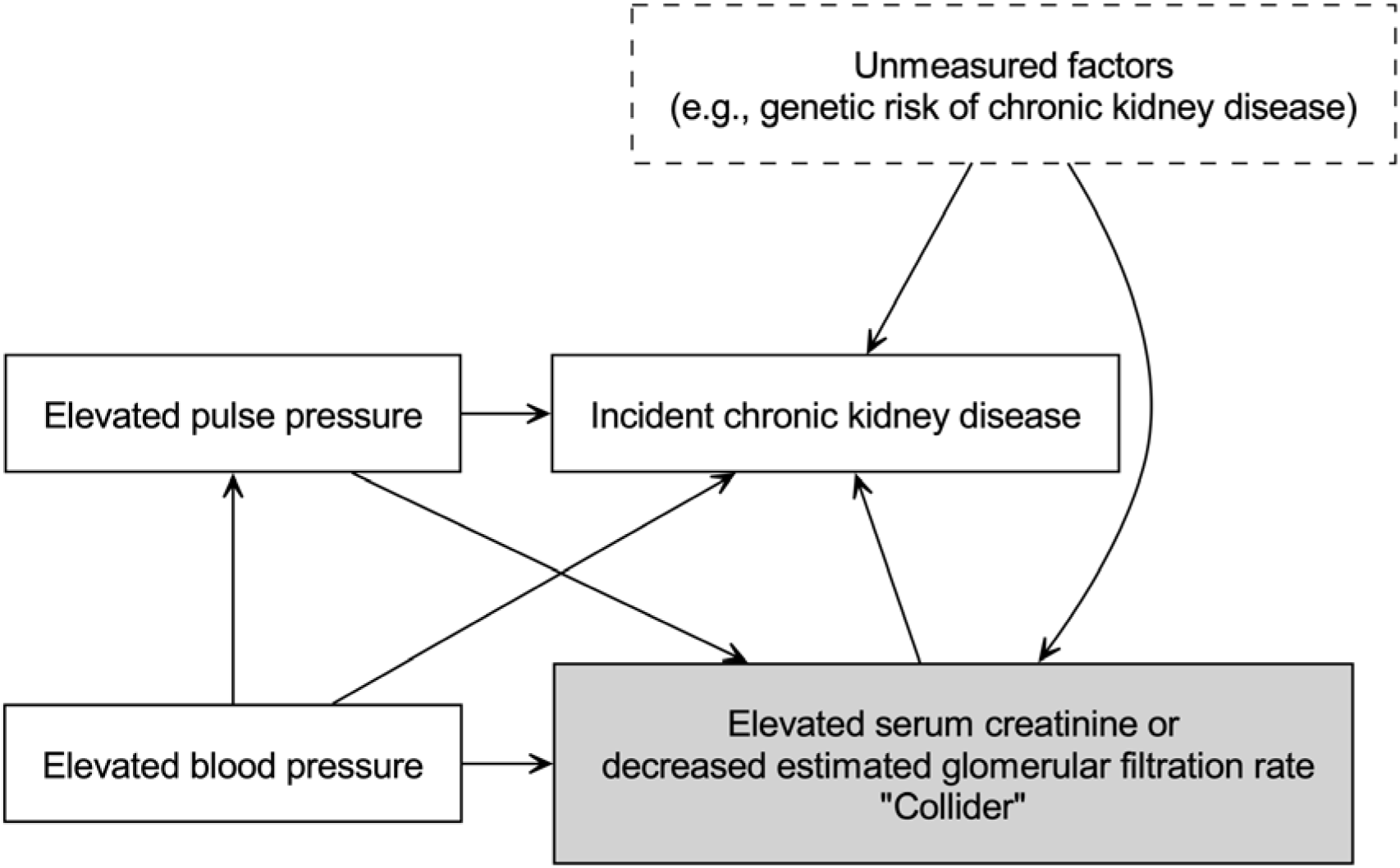
Causal directed acyclic graph used in the present study.

A two-tailed p-value of <0.05 was regarded to indicate statistical significance. The causal directed acyclic graph and flowchart were created using Python 3.11.11[40]. All other statistical analyses were conducted using Stata/MP 16.1 (StataCorp, College Station, TX, USA). This research followed the Strengthening the Reporting of Observational Studies in Epidemiology (STROBE) reporting guideline[41].

### Sensitivity analyses

The study included two sensitivity analyses. First, to examine the presence of sex differences in the relationship between time-varying PP and incident CKD, all analyses were further stratified by sex. Second, to address the limitation of eGFR in underestimating CKD diagnosis, CKD was defined as eGFR <60 mL/min/1.73 m^2^ and/or dipstick proteinuria ≥1+[42].

### Ethic

The data were anonymized before receipt. The Ethics Committee of Okayama University Graduate School of Medicine, Dentistry and Pharmaceutical Sciences and Okayama University Hospital approved this study (No. K1708-040). The ethics committee waived the need for informed consent. This research followed the Declaration of Helsinki and Japanese Ethical Guidelines for Medical and Biological Research involving Human Subjects.

## Results

Initially, 15788 participants (men: 41.0%; mean age at study entry: 62.3 years) were enrolled in the study. After applying the exclusion criteria (**Figure 2**), 8881 participants (men: 42.7%; mean age at study entry: 59.9 years; hypertensive status at study entry: 54.2%) remained in the final cohort, with checkup dates ranging from 6 April 1998 to 10 June 2024. At study entry, the distributions of the PP categories of <40, 40–<60, and ≥60 mmHg were 16.5%, 55.3%, and 28.2% for the total population, 27.9%, 68.1%, and 4.0% for non-hypertensive participants, and 6.8%, 44.5%, and 48.7% for hypertensive participants, respectively.

**Figure 2.**
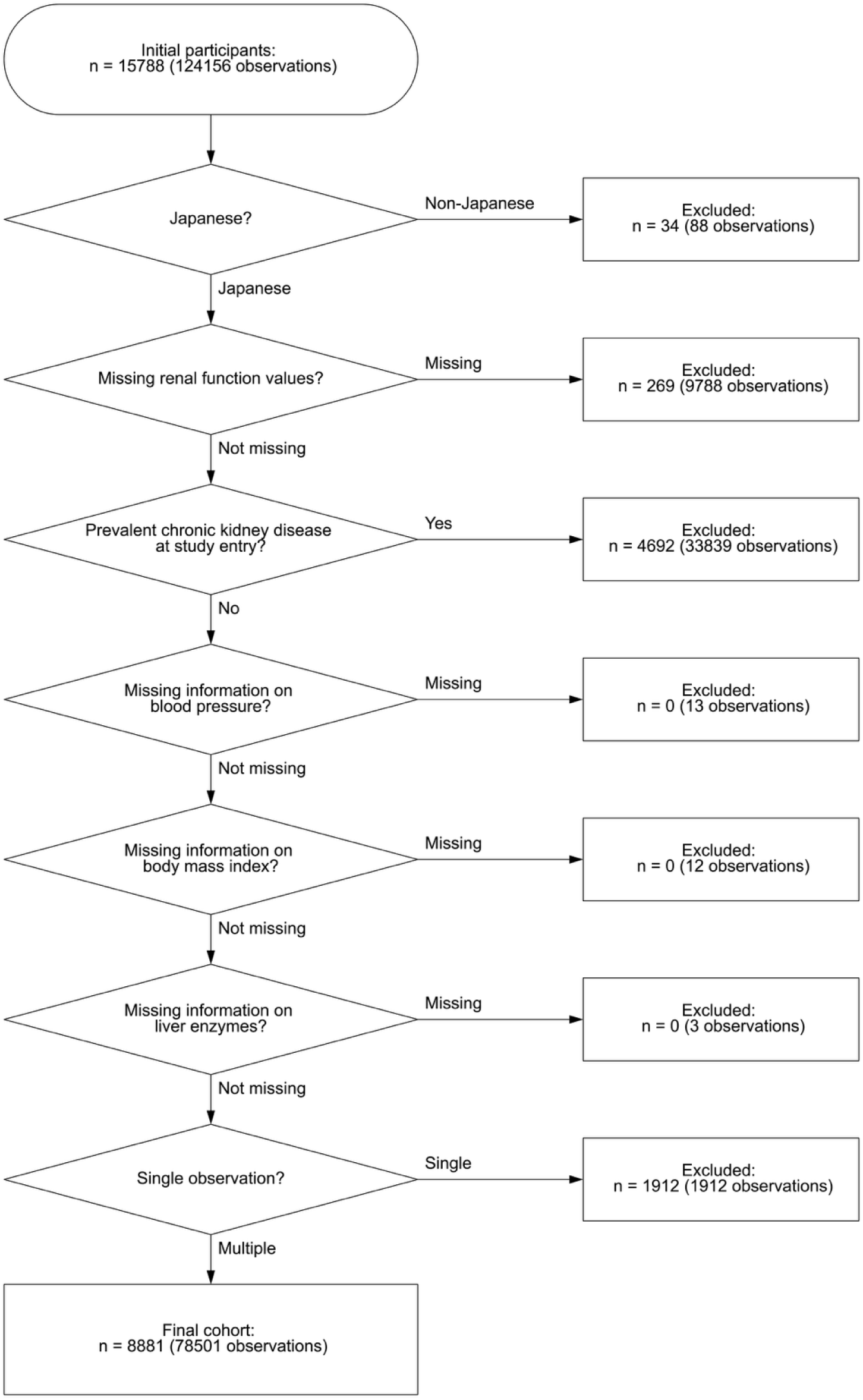
Flow diagram of the study participants.

The total follow-up times were 37922.1 person-years for non-hypertensive participants and 41909.8 person-years for hypertensive participants (**Tables 1 and 2**). During the entire follow-up period, the distributions of the PP categories of <40, 40–<60, and ≥60 mmHg were 25.2%, 69.1%, and 5.8% for non-hypertensive participants and 6.2%, 39.9%, and 53.9% for hypertensive participants, respectively. Because all variables were treated as time-varying variables in this analysis, some participants were classified into both the hypertensive and non-hypertensive groups due to changes in BP during follow-up. Regardless of the hypertension status, a higher time-varying PP was associated with a higher IR of CKD.

**Table 1.**
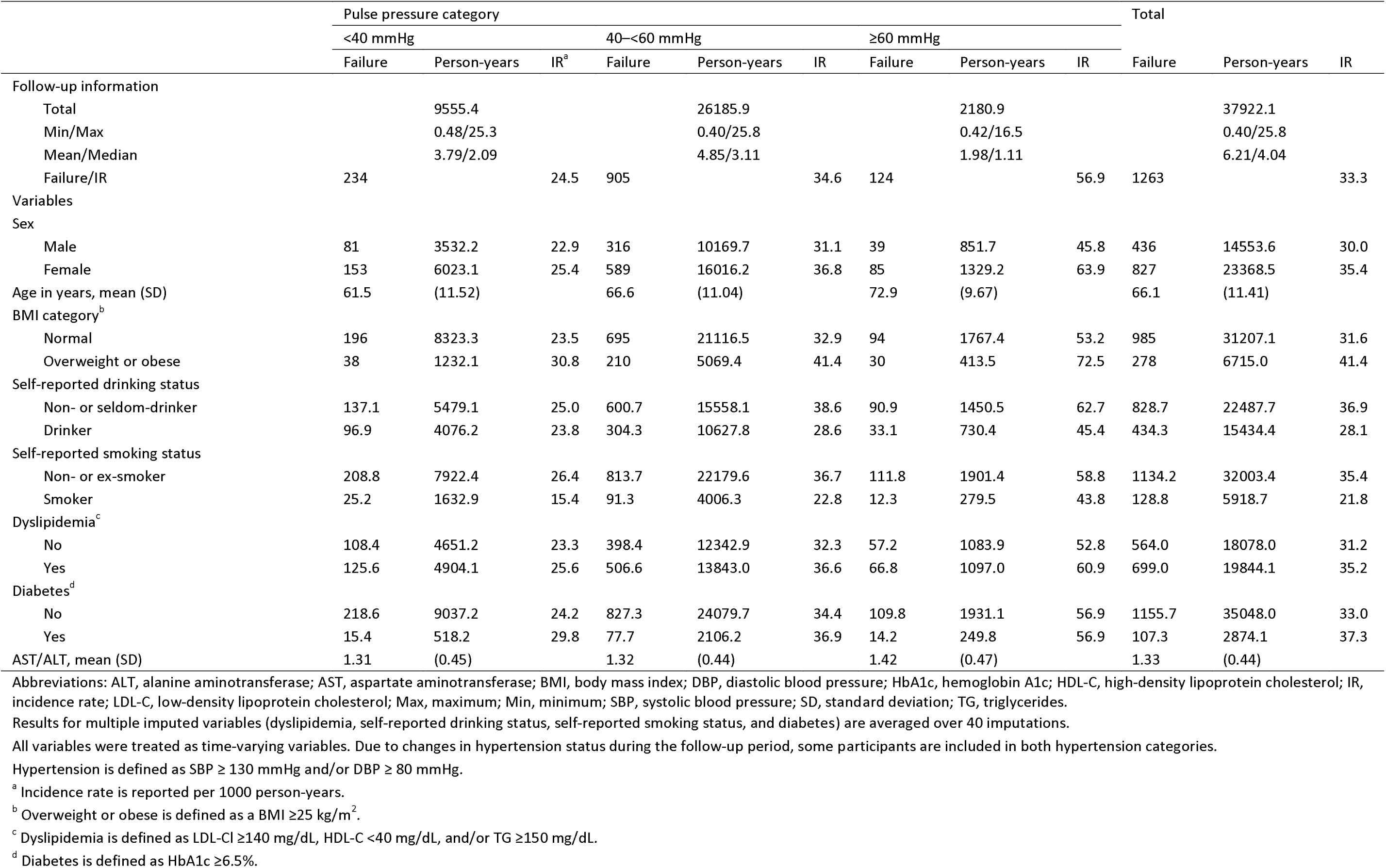
Descriptive statistics of all observations stratified by the time-varying pulse pressure category among 6106 non-hypertensive Japanese citizens of Zentsuji City (24662 observations, 1998–2024).

**Table 2.**
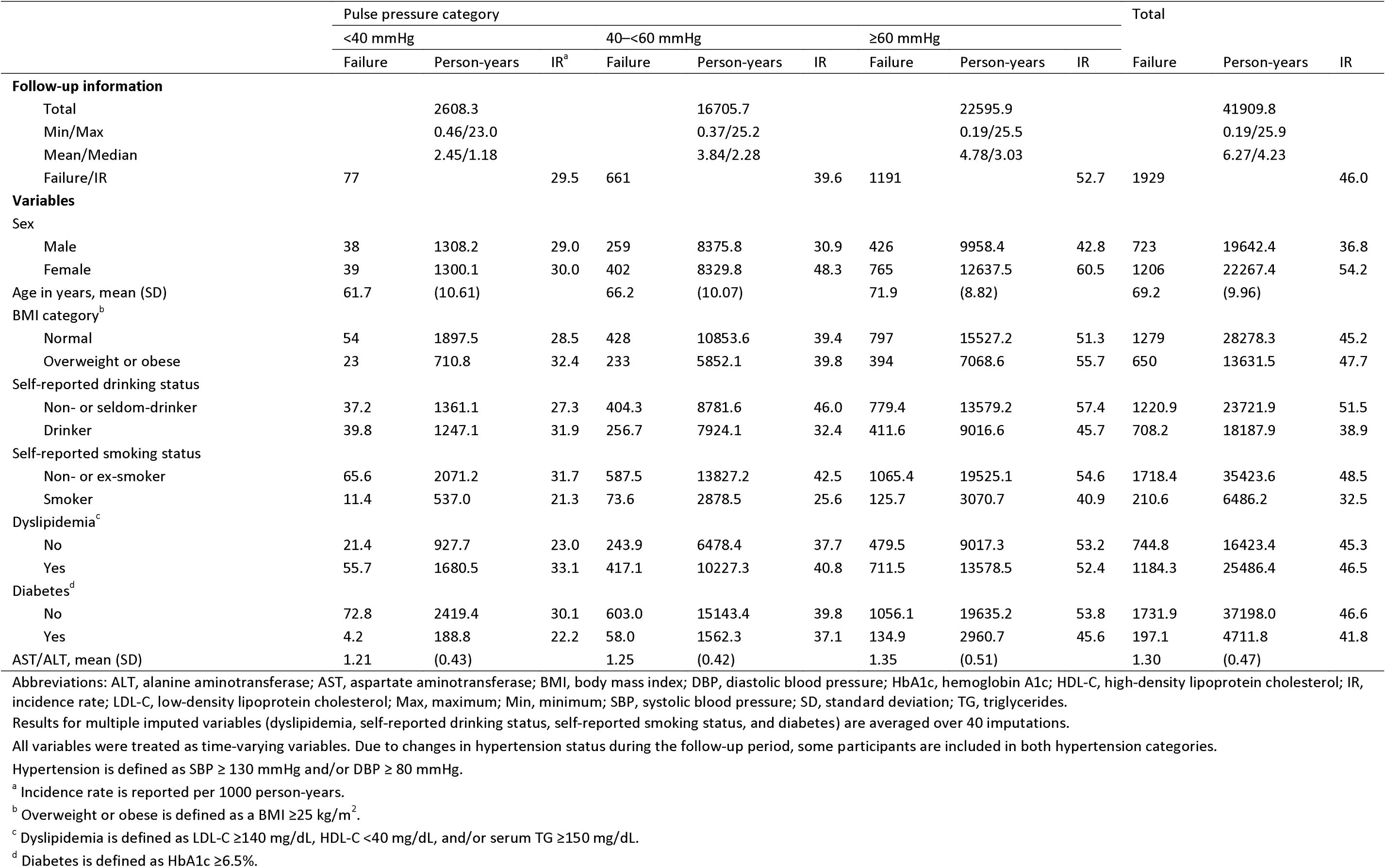
Descriptive statistics of all observations stratified by the time-varying pulse pressure category among 6682 hypertensive Japanese citizens of Zentsuji City (27564 observations, 1998–2024).

**Figure 3** presents the Kaplan–Meier curves for the PP categories stratified by hypertension status. Compared with PP <40 mmHg, higher PP showed a shorter survival time to CKD in participants with and without hypertension, based on point estimates (**Table 3**). A more distinct dose–response relationship was observed in non-hypertensive participants compared with hypertensive participants. In the highest category of time-varying PP ≥60 mmHg, the adjusted time ratios were 0.90 (95% confidence interval [CI]: 0.84–0.97) for non-hypertensive participants and 0.97 (95% CI: 0.93–1.01) for hypertensive participants, compared with the reference category of PP <40 mmHg. Hypertensive participants with higher time-varying PP categories had a 2%–3% shorter survival time to CKD by point estimate in the full model, with the 95% CIs crossing one.

**Figure 3.**
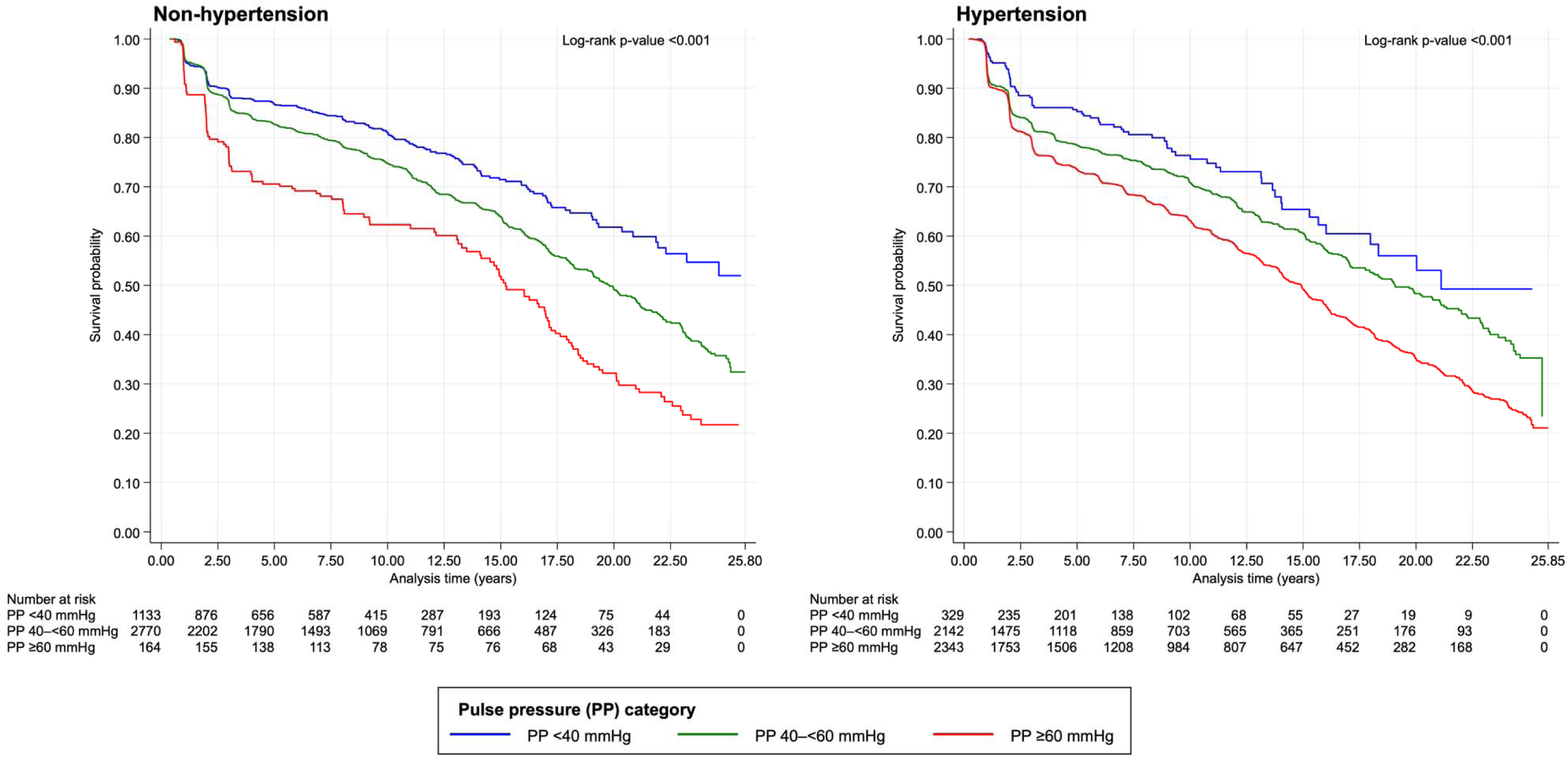
Kaplan–Meier curves stratified by pulse pressure category and hypertension status.

**Table 3.** New onset of chronic kidney disease according to the time-varying pulse pressure categories among 8881 Japanese citizens of Zentsuji City (Total time at risk: 79831.9 PY, 1998–2024) Non-hypertensive (Total time at risk: 37922.1 PY) Hypertensive (Total time at risk: 41909.8 PY)

| Pulse pressure classification | Non-hypertensive (Total time at risk: 37922.1 PY) |  |  |  |  |  | Hypertensive (Total time at risk: 41909.8 PY) |  |  |  |  |  |
| --- | --- | --- | --- | --- | --- | --- | --- | --- | --- | --- | --- | --- |
|  | Follow-up information |  |  | Models |  |  | Follow-up information |  |  | Models |  |  |
|  | Total PY | Failure | IR | Crude<br>TR (95% CI) | Model 1<br>aTR (95% CI) | Model 2<br>aTR (95% CI) | Total PY | Failure | IR | Crude<br>TR (95% CI) | Model 1<br>aTR (95% CI) | Model 2<br>aTR (95% CI) |
| <40 mmHg (reference) | 9555.4 | 234 | 24.5 | 1.00 | 1.00 | 1.00 | 2608.3 | 77 | 29.5 | 1.00 | 1.00 | 1.00 |
| 40–<60 mmHg | 26185.9 | 905 | 34.6 | 0.95<br>(0.91–0.99) | 0.96<br>(0.92–1.00) | 0.96<br>(0.92–1.00) | 16705.7 | 661 | 39.6 | 0.97<br>(0.91–1.03) | 0.98<br>(0.94–1.02) | 0.98<br>(0.94–1.02) |
| ≥60 mmHg | 2180.9 | 124 | 56.9 | 0.88<br>(0.82–0.95) | 0.90<br>(0.84–0.97) | 0.90<br>(0.84–0.97) | 22595.9 | 1191 | 52.7 | 0.95<br>(0.89–1.01) | 0.97<br>(0.93–1.02) | 0.97<br>(0.93–1.01) |

Abbreviations: ALT, alanine aminotransferase; AST, aspartate aminotransferase; aTR, adjusted time ratio; BMI, body mass index; CI, confidence interval; DBP, diastolic blood pressure; HbA1c, hemoglobin A1c; HDL-C, high-density lipoprotein cholesterol; IR, incidence rate; LDL-C, low-density lipoprotein cholesterol; PY, person-years; SBP, systolic blood pressure; TG, triglycerides; TR, time ratio.
All variables were treated as time-varying variables. Due to changes in hypertension status during the follow-up period, some participants are included in both hypertension categories.
Pulse pressure is calculated by subtracting DBP from SBP.
Hypertension is defined as SBP ≥130 mmHg and/or DBP ≥80 mmHg.
Overweight or obesity is defined as a BMI ≥25 kg/m<sup>2</sup>.
Dyslipidemia is defined as LDL-C ≥140 mg/dL, HDL-C <40 mg/dL, and/or TG ≥150 mg/dL.
Diabetes is defined as HbA1c ≥6.5%.
Model 1: Adjusted for sex (men [reference]/women) and age.
Model 2: Adjusted for both variables of Model 1, overweight or obesity (no [reference]/yes), self-reported alcohol intake (non- or seldom-drinker [reference]/drinker), self-reported smoking status (non- or ex-smoker [reference]/smoker), dyslipidemia (no [reference]/yes), diabetes (no [reference]/yes), and AST/ALT ratio.

## Sensitivity analyses

### First sensitivity analysis: Sex stratification

**Table S1** shows the results of the sensitivity analysis stratified by sex. In men, the distributions of the time-varying PP categories of <40, 40–60, and ≥60 mmHg were 24.3%, 69.9%, and 5.9% for non-hypertensive participants and 6.7%, 42.6%, and 50.7% for hypertensive participants, respectively. Among women, the distributions were 25.8%, 68.5%, and 5.7% for non-hypertensive participants and 5.8%, 37.4%, and 56.8% for hypertensive participants, respectively.

The results showed a somewhat similar trend in the main analysis; increased time-varying PP was associated with higher IR of CKD, regardless of sex and hypertension status (**Table 2**). However, women exhibited a clearer dose–response relationship between time-varying PP and incident CKD than men. Specifically, in the non-hypertensive group, women with time-varying PP ≥60 mmHg had up to 32% shorter survival time to CKD by point estimate, whereas men in the same category had a reduction of only 2%.

### Second sensitivity analysis: CKD definition using eGFR and dipstick proteinuria

**Table S2** shows the results for the sensitivity analysis in which CKD was regarded as eGFR <60 mL/min/1.73 m^2^ and/or proteinuria ≥1+. After exclusion, 8471 participants remained in the final cohort. When the CKD definition included eGFR and dipstick proteinuria, the total follow-up time was reduced by 7.7% for non-hypertensive participants and 12.2% for hypertensive participants. At study entry, the distributions of the PP categories of <40, 40–<60, and ≥60 mmHg were 16.6%, 55.7%, and 27.7% for the total population, 27.8%, 68.2%, and 4.0% for non-hypertensive participants, and 6.8%, 44.9%, and 48.3% for hypertensive participants, respectively. The distributions of the time-varying PP categories were similar to the main analysis at 25.3%, 69.2%, and 5.5% for non-hypertensive participants and 6.3%, 40.5%, and 53.2% for hypertensive participants, respectively (**Tables 1 and 2**).

In all PP categories, the IRs of CKD were higher than those in the main analysis. The estimation results were consistent with those in the main analysis for participants with and without hypertension, with the shortest time to CKD onset observed in non-hypertensive participants with PP ≥60 mmHg, which was 13% shorter (95% CI: 4%–21%) compared to those with PP <40 mmHg (**Table 3**).

## Discussion

This longitudinal study confirmed that elevated time-varying PP shortens the time to incident CKD, particularly in non-hypertensive participants. A dose–response relationship was observed, with a 10% reduction (95% CI: 3%– 16%) at maximum in non-hypertensive participants with time-varying PP ≥60 mmHg compared with PP <40 mmHg. The results for hypertensive participants showed only a 3% reduction with time-varying PP ≥60 mmHg, with the 95% CI crossing the null value. The results for non-hypertensive participants in this study were consistent with those for the Chinese non-hypertensive participants in the China Health and Retirement Longitudinal Study, with 1.34 times (95% CI: 1.02–1.75) higher risk of renal decline in those with baseline PP ≥60 mmHg compared with PP <60 mmHg[9].

There are three potential reasons why the results were clearer in non-hypertensive participants. First, the non-hypertensive participants with higher PP, a surrogate marker for arterial stiffness, may have been pre-hypertensive[1]. Healthy elastic arteries stretch during systole and recoil during diastole, ensuring efficient function that results in increased diastolic flow and lower SBP and PP. Conversely, arteries with severe rigidity do not stretch and the entire blood volume flows directly through them, leading to elevated SBP and PP and reduced diastolic flow, resulting in a strong association between higher BP and higher PP[43]. The trend of the time-varying PP distributions was reversed depending on the hypertension status in this study; the proportions of the follow-up time for time-varying PP ≥60 mmHg were 5.8% for non-hypertensive participants and 53.9% for hypertensive participants (**Tables 1 and 2**). Therefore, non-hypertensive participants with the highest PP category may have had normal BP; however, they may have been pre-hypertensive and could have begun to lose arterial flexibility.

Second, hypertensive participants may have a detection problem. As described above, participants with higher BP tended to have higher PP[43]. In hypertensive participants, only 6.2% of the entire follow-up time was in the reference category (<40 mmHg). The small number of hypertensive participants in the reference PP category may have reduced the statistical power, leading to wider CIs and less precise estimates. On the contrary, the reference PP category in non-hypertensive participants accounted for 25.2% of the total follow-up time, suggesting that these results were more robust than those for hypertensive participants.

Third, the missing information on antihypertensive medication use may have affected the results. As described in the Methods section, more than half of the self-reported medication data were missing due to a system-related issue, mainly among participants aged ≥75 years[19]. This missingness may have led to the misclassification of hypertensive participants on medication as non-hypertensive participants, leading to overestimation. However, according to the National Database of Health Insurance Claims and Specific Health Checkups of Japan, only 22.9% of all checkup participants aged 40–74 years in Kagawa Prefecture reported taking any antihypertensive medications in FY2022[44]. Moreover, when first-line antihypertensive medications (e.g., calcium channel blockers) are taken, they widen the arteries, which lowers BP and also reduces PP[45]. The proportion of hypertensive participants on antihypertensive medication may not have been large, and “true” hypertensive participants with well-controlled BP by medication were more likely to have been classified into the lower PP category, leading to underestimation[10]. This suggests that the missingness of antihypertensive medication may not have had a significant impact on the results.

We performed a further stratification by sex (**Table S1**). The trends were generally similar to those in the main analysis (**Table 3**). The study revealed that higher time-varying PP harms renal function faster in women than in men by point estimate, regardless of hypertension status. On the contrary, the association of time-varying PP and CKD onset was nearly null in men. This finding is consistent with the results in the Kansai Healthcare Study for male Japanese participants without adjustment for antihypertensive medication use, giving adjusted hazard ratios (95% CIs) of 0.84 (0.64–1.10) for PP of 41–<48 mmHg, 0.93 (0.71–1.21) for PP of 48–<56 mmHg, and 1.05 (0.81–1.35) for baseline PP ≥56 mmHg, compared with the reference category of <40 mmHg[8].

The more noticeable results in women may arise from their vulnerability to increased PP due to their smaller aortic diameter, which leads to higher PP, and limited aortic remodeling after midlife, which causes arteriosclerosis[1]. This may explain why the impact of elevated PP on renal function was greater in women than in men in the present study.

### Strengths and Limitations

The strengths of this research lie in the reliability of the data derived from long-term longitudinal administrative checkup records (1998–2024) managed by the local government in accordance with the protocol set by the MHLW[13,19]. Since the checkups target relatively healthy residents living at home and capture people’s conditions before severe disease progression, the findings of this study can be used for public health initiatives aimed at preventing CKD development through population-level interventions. Furthermore, the study treated the PP as a time-varying variable. Therefore, the study can more accurately reflect how long-term changes in PP impact the incidence of CKD.

However, several limitations of the study must be acknowledged. First, owing to the nature of the regular checkups implemented at a relatively low cost, data for carotid-femoral pulse wave velocity, as the gold standard test for aortic stiffness but a costly test in Japan, were unavailable for the study[46].

Second, the BP measurement method may not consistently reflect the average of two readings, as recommended by the MHLW[19,21]. As described in the Methods section, a previous report indicated that examinees with elevated BP were more likely to undergo a second measurement[21]. This introduces measurement bias because the selective remeasurement leads to differential accuracy depending on BP: participants with higher BP are more likely to have accurate results based on two readings, whereas those with lower BP often rely on a single reading, which is more susceptible to error. However, the direction of this bias remains uncertain because participants with elevated BP tended to undergo a second measurement regardless of their true hypertension status, potentially aiding more accurate classification.

Third, the vulnerability of eGFR may have influenced the results[47]. To understand this impact, as well as using the estimation equation for Japanese people when calculating the eGFR, we defined CKD using eGFR plus dipstick proteinuria ≥1+, and confirmed the robustness of the results (**Table S2**)[26,42].

Fourth, the study is subject to influence from selection bias, as the participants tended to be healthier and have higher socioeconomic status than the general population. This may make it difficult to generalize the results to other populations including higher proportions of unhealthy people[48,49]. Furthermore, the long follow-up period may lead to a built-in selection bias, as participants less likely to develop CKD remain in the study cohort, potentially leading to underestimation of the true association in the study[50].

Finally, residual bias from unmeasured confounders remains a limitation of this study (e.g., physical activity, protein intake, uric acid, depression, socioeconomic status)[51–55]. For example, physical activity is known to be associated with lower PP and has been shown to protect against the development of CKD[51,56]. The absence of physical activity data in the present study may therefore lead to an overestimation of the observed association. However, the direction and magnitude of the bias introduced by multiple unmeasured confounders cannot be determined.

## Conclusions

This longitudinal study in middle-aged and older Japanese subjects found that elevated time-varying PP ≥60 mmHg was associated with a 10% shorter time to CKD onset in non-hypertensive participants, but had attenuated results in hypertensive participants. These findings will be beneficial for the public health sectors when conducting a population-based health approach to CKD prevention in relatively healthy Asian adults without hypertension in the future.

## Data Availability

All data generated or analyzed during this study are included in this published article and its supplementary information files.

## Acknowledgments

We thank all participants in this study, Ayaka Nakatsu, Masako Matsumoto, Mayumi Kitadani, and the local government officers of Zentsuji City who kindly supported our research. We thank Alison Sherwin, PhD, and Angela Morben, DVM, ELS, from Edanz (https://jp.edanz.com/ac) for editing a draft of this manuscript.

## Author Contributions

Conceptualization, Y.O.; methodology, T.M., Y.O.; formal analysis, Y.O.; investigation, Y.O.; data curation, Y.O.; writing—original draft preparation, Y.O.; writing—review and editing, T.M., Y.O.; visualization, Y.O.; supervision, T.M.; project administration, Y.O.

