## Supplementary Tables for "Elevated pulse pressure and risk of chronic kidney disease by hypertension status: A longitudinal study in Japanese adults"

**Table S1**. New onset of chronic kidney disease according to the pulse pressure categories among 8881 Japanese citizens of Zentsuji City (Total time at risk: 79831.9 PY, 1998–2024)

| **Men** | **Non-hypertensive (Total time at risk: 14553.6 PY)** | | | | | | **Hypertensive (Total time at risk: 19642.4 PY)** | | | | | |
|---|---|---|---|---|---|---|---|---|---|---|---|---|
| | Follow-up information | | | Models | | | Follow-up information | | | Models | | |
| | | | | Crude | Model 1 | Model 2 | | | | Crude | Model 1 | Model 2 |
| Pulse pressure classification | Total PY | Failure | IR | TR (95% CI) | aTR (95% CI) | aTR (95% CI) | Total PY | Failure | IR | TR (95% CI) | aTR (95% CI) | aTR (95% CI) |
| <40 mmHg (reference) | 3532.2 | 81 | 22.9 | 1.00 | 1.00 | 1.00 | 1308.2 | 38 | 29.0 | 1.00 | 1.00 | 1.00 |
| 40–<60 mmHg | 10169.7 | 316 | 31.1 | 0.99 (0.95–1.03) | 0.99 (0.97–1.01) | 0.99 (0.97–1.01) | 8375.8 | 259 | 30.9 | 1.03 (0.97–1.10) | 1.01 (0.98–1.05) | 1.02 (0.98–1.05) |
| ≥60 mmHg | 851.7 | 39 | 45.8 | 0.98 (0.92–1.05) | 0.99 (0.95–1.02) | 0.98 (0.95–1.02) | 9958.4 | 426 | 42.8 | 1.03 (0.97–1.10) | 1.01 (0.97–1.04) | 1.01 (0.97–1.04) |
| **Women** | **Non-hypertensive (Total time at risk: 23368.5 PY)** | | | | | | **Hypertensive (Total time at risk: 22267.4 PY)** | | | | | |
| | Follow-up information | | | Models | | | Follow-up information | | | Models | | |
| | | | | Crude | Model 1 | Model 2 | | | | Crude | Model 1 | Model 2 |
| Pulse pressure classification | Total PY | Failure | IR | TR (95% CI) | aTR (95% CI) | aTR (95% CI) | Total PY | Failure | IR | TR (95% CI) | aTR (95% CI) | aTR (95% CI) |
| <40 mmHg (reference) | 6023.1 | 153 | 25.4 | 1.00 | 1.00 | 1.00 | 1300.1 | 39 | 30.0 | 1.00 | 1.00 | 1.00 |
| 40–<60 mmHg | 16016.2 | 589 | 36.8 | 0.94 (0.88–0.99) | 0.85 (0.67–1.09) | 0.86 (0.67–1.09) | 8329.8 | 402 | 48.3 | 0.91 (0.82–1.01) | 0.90 (0.79–1.02) | 0.88 (0.76–1.01) |
| ≥60 mmHg | 1329.2 | 85 | 63.9 | 0.84 (0.76–0.93) | 0.67 (0.40–1.14) | 0.68 (0.41–1.14) | 12637.5 | 765 | 60.5 | 0.91 (0.82–1.01) | 0.89 (0.78–1.02) | 0.87 (0.75–1.01) |

Abbreviations: ALT, alanine aminotransferase; AST, aspartate aminotransferase; aTR, adjusted time ratio; BMI, body mass index; CI, confidence interval; DBP, diastolic blood pressure; HbA1c, hemoglobin A1c; HDL-C, high-density lipoprotein cholesterol; IR, incidence rate; LDL-C, low-density lipoprotein cholesterol; PY, person-years; SBP, systolic blood pressure; TG, triglycerides; TR, time ratio.

All variables were treated as time-varying variables. Due to changes in hypertension status during the follow-up period, some participants are included in both hypertension categories.

Pulse pressure is calculated by subtracting diastolic BP from systolic BP.

Hypertension is defined as SBP ≥130 mmHg and/or DBP ≥80 mmHg.

Overweight or obesity is defined as a BMI ≥25 kg/m$^2$.

Dyslipidemia is defined as LDL-C ≥140 mg/dL, HDL-C <40 mg/dL, and/or TG ≥150 mg/dL.

Diabetes is defined as HbA1c ≥6.5%.

Model 1: Adjusted for age.

Model 2: Adjusted for age, overweight or obesity (no [reference]/yes), self-reported alcohol intake (non- or seldom-drinker [reference]/drinker), self-reported smoking status (non- or ex-smoker [reference]/smoker), dyslipidemia (no [reference]/yes), diabetes (no [reference]/yes), and AST/ALT ratio.

**Table S2**. New onset of chronic kidney disease according to the pulse pressure categories among 8471 Japanese citizens of Zentsuji City, CKD defined as eGFR <60 mL/min/1.73m² and/or proteinuria (Total time at risk: 71784.9 PY, 1998–2024)

| | **Non-hypertensive (Total time at risk: 34985.7 PY)** | | | | | | **Hypertensive (Total time at risk: 36799.2 PY)** | | | | | |
|---|---|---|---|---|---|---|---|---|---|---|---|---|
| | Follow-up information | | | Models | | | Follow-up information | | | Models | | |
| | | | | Crude | Model 1 | Model 2 | | | | Crude | Model 1 | Model 2 |
| Pulse pressure classification | Total PY | Failure | IR | TR (95% CI) | aTR (95% CI) | aTR (95% CI) | Total PY | Failure | IR | TR (95% CI) | aTR (95% CI) | aTR (95% CI) |
| <40 mmHg (reference) | 8864.4 | 282 | 31.8 | 1.00 | 1.00 | 1.00 | 2315.1 | 97 | 41.9 | 1.00 | 1.00 | 1.00 |
| 40–<60 mmHg | 24198.8 | 1055 | 43.6 | 0.95 (0.91–0.99) | 0.94 (0.89–0.99) | 0.95 (0.90–1.00) | 14918.4 | 780 | 52.3 | 0.97 (0.91–1.04) | 0.98 (0.93–1.03) | 0.98 (0.93–1.03) |
| ≥60 mmHg | 1922.6 | 132 | 68.7 | 0.87 (0.81–0.94) | 0.87 (0.79–0.96) | 0.87 (0.79–0.96) | 19565.7 | 1369 | 70.0 | 0.94 (0.88–1.01) | 0.96 (0.92–1.01) | 0.96 (0.91–1.01) |

Abbreviations: ALT, alanine aminotransferase; AST, aspartate aminotransferase; aTR, adjusted time ratio; BMI, body mass index; CI, confidence interval; DBP, diastolic blood pressure; HDL-C, HbA1c, hemoglobin A1c; HDL-C, high-density lipoprotein cholesterol; IR, incidence rate; LDL-C, low-density lipoprotein cholesterol; PY, person-years; SBP, systolic blood pressure; TG, triglycerides; TR, time ratio.

All variables were treated as time-varying variables. Due to changes in hypertension status during the follow-up period, some participants are included in both hypertension categories.

Pulse pressure is calculated by subtracting diastolic BP from systolic BP.

Hypertension is defined as SBP ≥130 mmHg and/or DBP ≥80 mmHg.

Overweight or obesity is defined as a BMI ≥25 kg/m².

Dyslipidemia is defined as LDL-C ≥140 mg/dL, HDL-C <40 mg/dL, and/or TG ≥150 mg/dL.

Diabetes is defined as HbA1c ≥6.5%.

Model 1: Adjusted for sex (men [reference]/women) and age.

Model 2: Adjusted for both variables of Model 1, overweight or obesity (no [reference]/yes), self-reported alcohol intake (non- or seldom-drinker [reference]/drinker), self-reported smoking status (non- or ex-smoker [reference]/smoker), dyslipidemia (no [reference]/yes), diabetes (no [reference]/yes), and AST/ALT ratio.
